# Consumption of processed meat and its interactions with alcohol drinking and polygentic risk scores on breast cancer risk: a cohort study in the UK biobank

**DOI:** 10.1101/2022.08.30.22279400

**Authors:** Pingxiu Zhu, Yanyu Zhang, Shuqing Zou, Xingxing Yu, Mengjie Song, Moufeng Lin, Haomin Yang

## Abstract

**Background:** Processed meat and alcohol have been consistently associated with breast cancer risk, but evidence for their effects in women with different genetic susceptibility of breast cancer is scarce, and little is known about their interactions.

**Methods:** We analyzed data from 260,779 female participants in the UK Biobank. Multivariable adjusted Cox proportional hazards models were used to estimate hazard ratios (HR) and 95% confidence intervals (CI) for associations between processed meat and breast cancer risk. We further assessed its interaction with alcohol intake and polygenic risk score (PRS) for breast cancer.

**Results:** Processed meat intake more than once a week was positively associated with risk of breast cancer, especially in women took alcohol ≥1/d (HR=1.50, 95% CI=1.17-1.93), and in women who usually took alcohol together with meals (HR=1.70, 95% CI=1.21-2.39, P for interaction=0.048). The association between processed meat and breast cancer did not differ by menopausal status. When further stratified by PRS, processed meat more than once a week intake was associated with risk of breast cancer (HR=1.17, 95% CI=1.02-1.35) in women with the highest quantile of PRS, and additive interaction was found between them.

**Conclusions:** Processed meat was associated with risk of breast cancer in women, and the effect was stronger in those who took alcohol together with the meal and with high PRS of breast cancer, suggesting the focus of future preventive measures on these women.

**Funding:** This work was supported by the Natural Science Foundation of Fujian Province [grant no: 2021J01721], the Startup Fund for High-level Talents of Fujian Medical University [grant no: XRCZX2020007], Startup Fund for Scientific Research, Fujian Medical University [grant no: 2019QH1002] and Laboratory Construction Program of Fujian Medical University [grant no: 1100160208].

## Introduction

On a global scale, breast cancer is the most common cancer among women and the second leading cause of cancer death^[1]^. Given the international variations in breast cancer rates and trends^[2]^, it’s important to identify modifiable lifestyle risk factors to reduce the risk of breast cancer, such as physical activity, breast feeding and alcohol consumption. Processed meat consumption has been identified as a risk factor for breast cancer in several studies^[3]^, while there is less information on the effect among different subgroups of women. Identification of subgroups of women with higher risk of breast cancer associated with processed meat consumption could result to targeted intervention for women and increase the cost effectiveness of interventions.

Alcohol consumption has long been thought to play a major role in the development of breast cancer^[4]^. Some epidemiological studies have investigated the association between alcohol intake and breast cancer risk^[5,6]^. However, the interaction between processed meat and alcohol consumption has been shown in esophageal cancer^[7]^, while no study has assessed their synergistic effect on risk of breast cancer.

Besides the interaction with other life style factors on breast cancer risk, the association between processed meat and breast cancer could also be influenced by genetic factors. Recent GWAS studies has reveals 313 SNPs to be associated with breast cancer risk and could be used as a tool for risk stratification. However, it is still unclear whether the effect of processed meat differs according to different genetic predisposition to breast cancer, which is important for personalized prevention.

The aim of this study was to evaluate the association between processed meat intake and the risk of breast cancer, and to investigate whether the association was influenced by alcohol consumption, especially when the alcohol was taken with meals. We also examined the association between processed meat intake and breast cancer risk according to genetic susceptibility to breast cancer measured using polygenic risk score.

## Methods

### Study population, exposure and outcome

Between 2006 and 2010, 503 317 participants (women N= 273,382) consented to participate the baseline assessment of the UK Biobank study^[8]^. Participants were followed up from their date of enrollment until the date of diagnosis of breast cancer, date of withdrawal from the study, date of death, loss of follow up or until the end of follow-up (31 March 2016 for England and Wales, 31 October 2015 for Scotland), whichever came first. Information on breast cancer diagnosis, was obtained by using unique personal identification numbers to link the cohort to the National Health Service (NHS) Digital for England and Wales, and National Records of Scotland, NHS Central Register for Scotland. The ICD-10 code C50 was used to identify breast cancer diagnoses in the cancer register. Women with breast cancer before participating in the UK biobank were excluded from the analysis. The date of death was retrieved from death certificates held by the NHS Information Center and the NHS Central Register. The study was approved by The National Information Governance Board for Health and Social Care and the NHS North West Multicentre Research Ethics Committee (06/MRE08/65), and participants provided informed consent at baseline and to be followed up using data-linkage. The inclusion and exclusion criteria for the study populations are shown in supplementary figure 1.

### Diet group classification

Dietary intake data were collected at recruitment using a self-reported touchscreen questionnaire (http://biobank.ctsu.ox.ac.uk/showcase/showcase/docs/TouchscreenQuestionsMainFinal.pdf). Processed meats included bacon, ham, sausages, meat pies, kebabs, burgers and chicken nuggets. Frequency of consumption were coded into three categories (never, less than once a week and more than once a week). Participants with missing information or reported “prefer not to answer” or “do not know” were excluded from the analysis. Frequency of alcohol drinking was also collected in the questionnaire, and we dichotomized them into whether or not had daily alcohol drinking. We further included the question on whether the alcohol was usually taken with meal.

### Polygenic risk score

Blood samples from the participants were collected when they joined the cohort and were genotyped using the UK Biobank Axiom array. A brief description of the procedure for genotype calling, array design, sample handling, quality control, and imputation for the UK biobank samples were described elsewhere^[9]^. To assess whether the effect of processed meat intake differed according to genetic susceptibility to breast cancer, significant SNPs reported in a recent meta-analysis of breast cancer GWAS were selected to construct polygenic risk scores for breast cancer overall and by estrogen receptor (ER) status^[10]^. For all individuals, to calculate the weighted polygenic risk score (PRS) by the following formula:

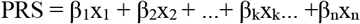

where β_k_ is the per-allele log odds ratio (OR) of breast cancer associated with SNP k, xk is the allele dose of the same SNP (0, 1, 2), and n is the total number of the breast cancer SNPs contained in the configuration file. The overall, ER+ and ER-PRS were respectively categorized into quartiles. Detailed information of PRS score generation is provided in the Supplementary Table 2.

### Statistical analysis

Cox proportional hazards models were used to assess the associations between processed meat intake and with breast cancer risk adjusting for age, smoking (never, previous, current), ethnicity (five groups where possible: White, Mixed other, Asian or Asian British, Black or Black British, and unkown), physical activity level (metabolic equivalents task units in quartiles), Townsend deprivation index (quintile), alcohol intake frequency (≥1/d, <1/d), employment status (in paid employment, pension, not in paid employment), educational qualifications (college or university degree/vocational qualification; national examination at ages 17-18 years; national examination at age 16 years; other qualifications were treated as missing), body mass index (BMI, <18.5, <25, <30, ≥30 kg/m^2^), 22 UKB centers, number of births (0, 1, 2, ≥ 3), age at menarche —years (<13, 13-15, >15 and <30), menopausal status (no, yes, not sure - had a hysterectomy, not sure - other reason), age at first birth—years (<23, 23-27, >27), ever use of oral contraceptive pill use (no, yes), ever use of hormone replacement therapy (no, yes), family history of breast cancer (no, yes). Missingness in the covariates were categorized into a separate category.

To test for multiplicative interaction between the processed meat intake and alcohol consumption, and the processed meat intake and PRS, an interaction term was included in the regression models and we tested the interaction using likelihood ratio (LR) test. Interaction in the additive scale for processed meat and PRS was estimated using relative excess risk due to interaction (RERI), and a bootstrap approach was used to estimate the p-values. We further stratified the analysis by alcohol intake frequency, alcohol taken with meal, quartile of PRS, and menopausal status.

All statistical analyses were performed using Stata 15.1. All P values were two-sided, and a P value of less than 0.05 was considered statistically significant.

## Results

### Baseline characteristics

Among all the 273,382 women in the UK Biobank, 54 were dropped and withdrew consent, 1,078 women did not have available data on processed meat intake and 11,471 women were excluded due to breast cancer diagnosis before baseline, resulting in 260,779 participants and 5,523 incident breast cancer cases in our study. The median follow-up time was 6.7 years and the incidence rate was 318.761/100000 person-year in the cohort.

Table 1 shows characteristics of all participants by categories of processed meat intake. There were 49% of participants consumed processed meat once or more weekly. Participants reporting higher processed meat intakes were more likely among those with smoking, White ethnicity, less physically activity, more affluent areas (measured by Townsend score), more alcohol drinking, paid employment, a university/college degree/NVQ, two or more children, postmenopausal status, age at first birth more than 23 years old, oral contraceptives pill, hormone replacement therapy, no family history of breast cancer, and a higher BMI.

**Table 1.**
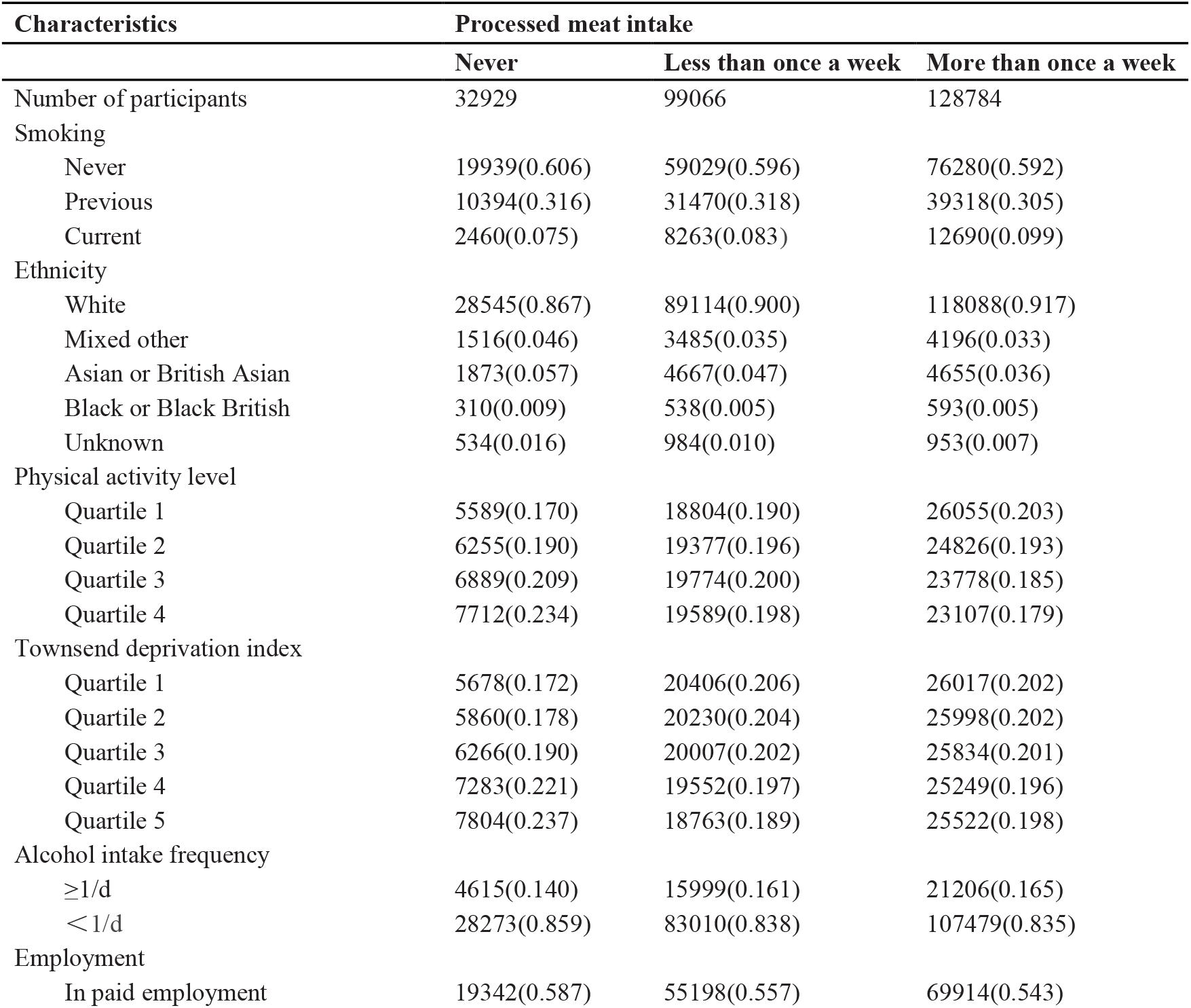
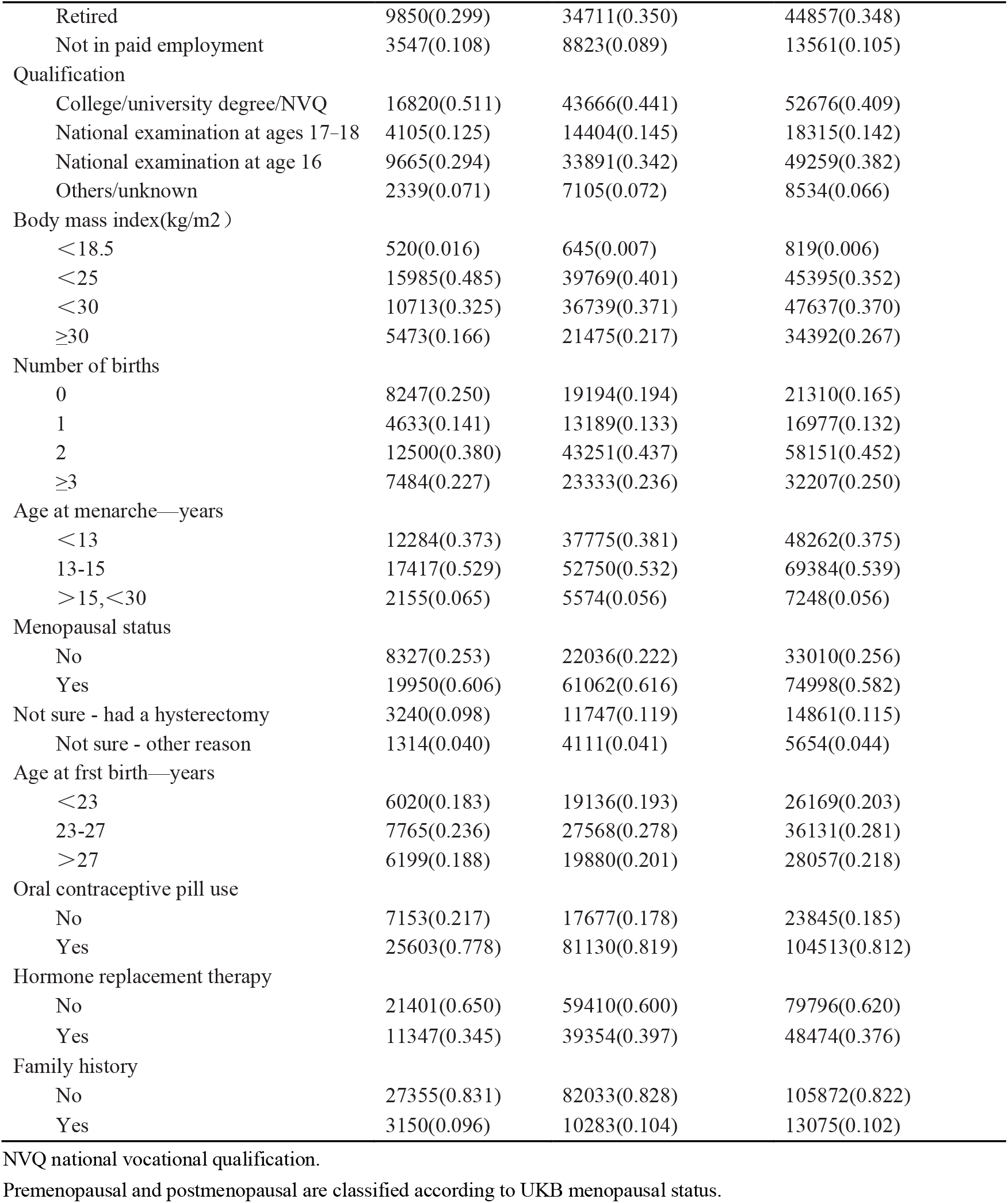
Baseline characteristics in the UK Biobank, by frequency of processed meat consumption (N= 260,779).

Processed meat intake was positively associated with risk of breast cancer, (HR more than once a week =1.13, 95% CI=1.04-1.24, Table 2). Among women took alcohol ≥ 1/d, processed meat intake was associated with a higher risk of breast cancer (HR=1.50, 95% CI=1.17-1.93), while we did not observe this association among women who took alcohol <1/d. When stratified the analysis by menopausal status, processed meat was associated with risk of breast cancer in postmenopausal women, especially in those took alcohol ≥1/d, while no association or interaction was observed in premenopausal women (Supplementary Table 1).

**Table 2.**
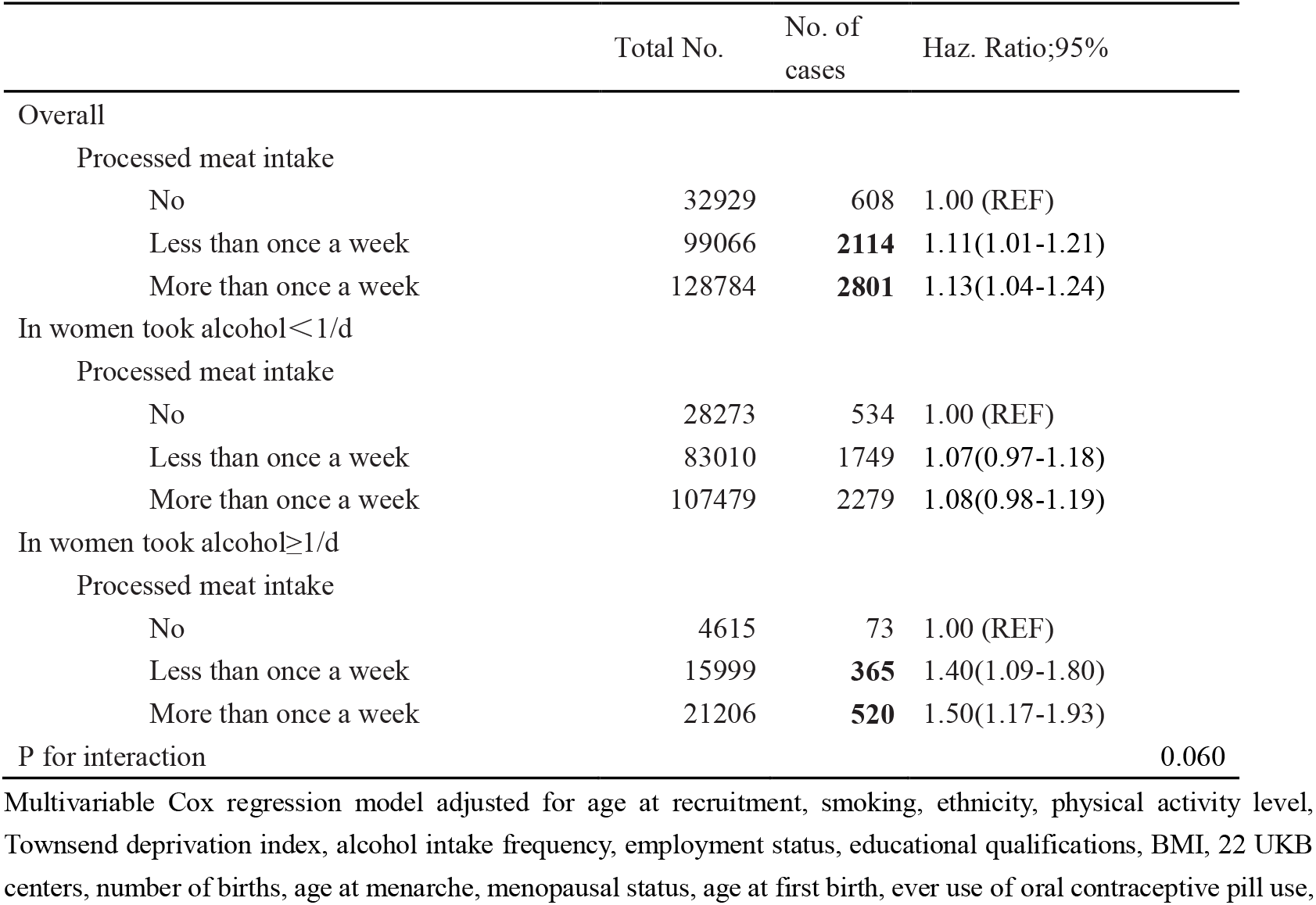
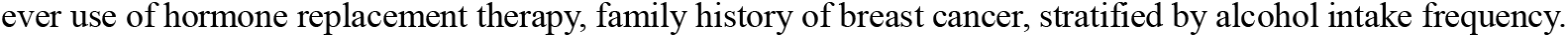
The association between processed meat and breast cancer risk, by frequency of alcohol consumption

For women who usually took alcohol together with the meal, processed meat intake more than once a week was associated with increased risk of breast cancer (HR=1.20, 95% CI=1.03-1.40), while the effect was attenuated in those who did not usually took alcohol with the meal. Furthermore, the interaction between processed meat and alcohol on breast cancer risk was only observed in those who usually took alcohol together with the meal (Table 3, P for interaction=0.048). In these women, processed meat intake was associated with 70% increased risk of breast cancer (HR=1.70, 95% CI=1.21-2.39).

**Table 3.** The interaction between processed meat and alcohol on breast cancer risk, by alcohol and meal

|  | Alcohol usually taken with meals<br>(Yes) |  |  | Alcohol usually taken with<br>meals (No) |  |  |
| --- | --- | --- | --- | --- | --- | --- |
|  | Total<br>No. | No. of<br>cases | Haz. Ratio;95% | Total<br>No. | No. of<br>cases | Haz. Ratio;95% |
| Overall |  |  |  |  |  |  |
| Processed meat intake |  |  |  |  |  |  |
| No | 11297 | 206 | 1.00 (REF) | 9846 | 175 | 1.00 (REF) |
| Less than once a week | 38670 | 830 | 1.13(0.97-1.32) | 33443 | 720 | 1.15(0.97-1.36) |
| More than once a week | 46121 | **1046** | 1.20(1.03-1.40) | 48816 | 1037 | 1.14(0.97-1.34) |
| In women took alcohol<1/d |  |  |  |  |  |  |
| Processed meat intake |  |  |  |  |  |  |
| No | 8733 | 168 | 1.00 (REF) | 7795 | 140 | 1.00 (REF) |
| Less than once a week | 29491 | 626 | 1.049(0.88-1.25) | 26625 | 559 | 1.12(0.93-1.35) |
| More than once a week | 34961 | 762 | 1.08(0.92-1.29) | 38775 | 801 | 1.12(0.93-1.34) |
| In women took alcohol≥1/d |  |  |  |  |  |  |
| Processed meat intake |  |  |  |  |  |  |
| No | 2564 | 38 | 1.00 (REF) | 2051 | 35 | 1.00 (REF) |
| Less than once a week | 9179 | **204** | 1.48(1.05-2.10) | 6818 | 161 | 1.28(0.88-1.84) |
| More than once a week | 11160 | **284** | 1.70(1.21-2.39) | 10041 | 236 | 1.25(0.87-1.79) |
| P for interaction |  |  | **0.048** |  |  | 0.397 |

Multivariable Cox regression model adjusted for age at recruitment, smoking, ethnicity, physical activity level, Townsend deprivation index, alcohol intake frequency, employment status, educational qualifications, BMI, 22 UKB centers, number of births, age at menarche, menopausal status, age at first birth, ever use of oral contraceptive pill use, ever use of hormone replacement therapy, family history of breast cancer, stratified by alcohol intake frequency, alcohol and meal.

When stratified by genetic susceptibility to breast cancer using PRS in quartiles, (Table 4). The only observed associations between processed meat intake and breast cancer risk were in women with highest quartile of overall PRS (HR=1.16, 95% CI = 1.01-1.34) and ER+ PRS (HR=1.18, 95% CI=1.02-1.36). The RERI of PRS, ER+ PRS and ER-PRS were0.437, 0.455 and 0.334, respectively. A statistical significant additive interaction between ER+ PRS and processed meat intake was observed in the highest quartile (p for RERI=0.034).

**Table 4.**
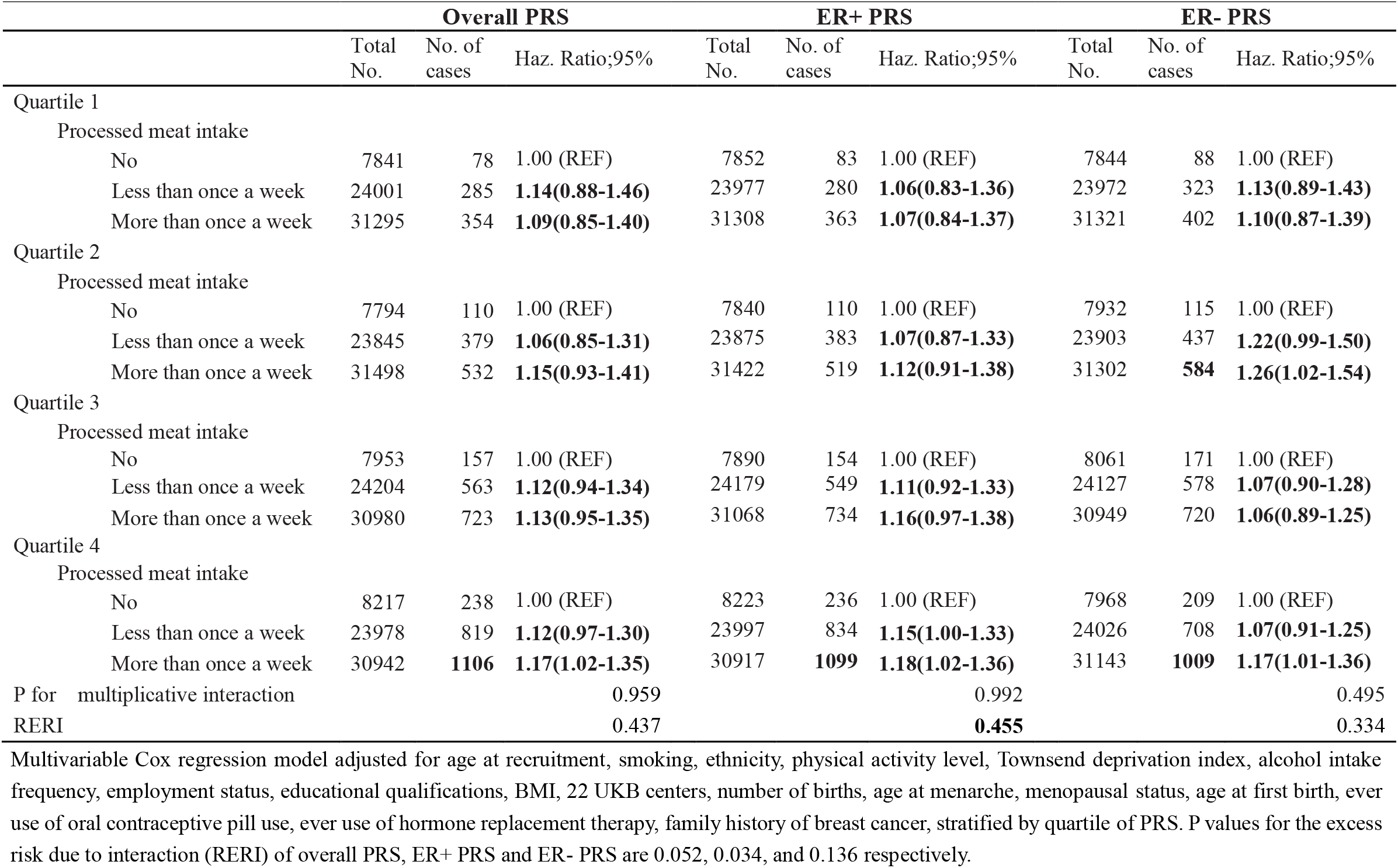
The interaction between processed meat and polygenic risk score on breast cancer risk

## Discussion

We observed a significant association between processed meat consumption and risk of breast cancer, especially in those women with high frequency of alcohol intake, and in women with alcohol usually taken with meals. We also found stronger association between processed meat and breast cancer in women with higher ER+ PRS.

Evidence from several previous prospective studies and meta-analysis supported these findings and has also shown an association between processed meat intake and breast cancer risk ^[11,12]^. Processed red meat was high in added nitrites/nitrates, amines and heme iron, while in animal and human biomonitoring studies, this combination has been shown to increase endogenous NOC formation^[13,14]^. Moreover, high-temperature cooking methods used in processed meat can result in the formation of compounds such as HCAs (including PhIP) and PAHs^[15,16]^ which have been associated with breast tumors in animal studies^[17-19]^ and human^[20-25]^. These compounds exert carcinogenic effects either through direct DNA damage (formation of DNA adducts) or through other mechanisms (such as the estrogenic properties of PhIP)^[26,27]^.

We found an interaction between processed meat intake and alcohol consumption on breast cancer risk. Recent large prospective studies found that alcohol intake was related to breast cancer risk^[28-30]^, which was probably hormonally driven^[31-33]^. Alcohol has also been suggested to have toxic effects and these effects were mediated by DNA damage and carcinogenic effects of alcohol and through mutagenesis by acetaldehyde and by induction of oxidative damage^[34-36]^. Simultaneous consumption of processed meat and alcohol might increase CYP2E1 expression, leading to increased oxidative stress and DNA damage. CYP2E1 then enhanced the activation of DNA damaged ROS by ethanol and ROS production, and activated PhIP through a single electron oxidation, which could trigger or maintain the tumor environment^[34]^. One plausible mechanisms for the synergic effect between processed meat and alcohol is that, when alcohol is consumed together with processed meat, alcohol may enhance the penetration of carcinogenic compounds (the genotoxicity of PhIP in the presence of ethanol) in processed meat as a solvent^[7]^. This possibility was further confirmed by our finding that the interaction was only observed among women who usually took alcohol together with the meal.

In the current study, we observed the association between processed meat intake and risk of breast cancer in women with the highest quartile of the PRSs. Moreover, the slight higher risk of breast cancer among women with high processed meat and ER+ PRS than overall PRS may reflect a stronger association with ER-positive disease^[39,40]^. Our finding of the additive interaction between ER+ PRS and processed meat further suggested the role of genetic testing in individualized dietary intervention for breast cancer. However, to date, fewer studies to our knowledge have examined the combined effect of processed meat and breast cancer associated genes on breast cancer risk. Further studies are need to confirm our findings.

The main strength of our study is having a large sample size and a population-based cohort design. Other strengths include the UK Biobank cohort containing rich lifestyle and genetic data, which allowed us to explore the gene and lifestyle interactions for the associations investigated. However, our study has several limitations. For one hand, there is no real ration of alcohol consumption, but by frequency of alcohol consumption. For another, the data from the touchscreen dietary questionnaire might suffer from recall bias. Finally, given the observational nature of this study, it is possible that there is still unmeasured confounding, or residual confounding in our analysis.

In conclusion, our findings support the view that processed meat can boost the risk of breast cancer, and it’s more obvious in postmenopausal women and women with genetic predisposition to breast cancer. Furthermore, the association between breast cancer and processed meat were stronger in women took alcohol, especially when the alcohol was usually taken with meals. A combined intervention by reducing alcohol consumption with meals or avoiding processed meats when taking alcohol might contribute to the prevention of breast cancer in women with genetically high risk.

## Data Availability

Data from the UK Biobank (http://www.ukbiobank.ac.uk/) are available to researchers upon application. This research was conducted using the UK Biobank Resource under Application 61083.

## Funding

This work was supported by the Natural Science Foundation of Fujian Province [grant no: 2021J01721], the Startup Fund for High-level Talents of Fujian Medical University [grant no: XRCZX2020007], Startup Fund for Scientific Research, Fujian Medical University [grant no: 2019QH1002] and Laboratory Construction Program of Fujian Medical University [grant no: 1100160208]. The funders had no role in the study design, data collection, analyses, data interpretation, writing the manuscript, or in the decision to submit the manuscript for publication.

## DECLARATIONS

### Competing interests

The authors declare no competing interest.

### Ethics approval and consent to participate

The UK Biobank was approved by the National Information Governance Board for Health and Social Care and the National Health Service North West Centre for Research Ethics Committee (Ref: 11/NW/0382, 17 June 2011). All participants gave informed consent to participate and be followed-up.

### Author contributions

PZ, YZ and HY had full access to all data, and take responsibility for the integrity of the data and the accuracy of the analysis. HY and ML conceived and designed the study. All authors acquired, analyzed, or interpreted the data. PZ and HY drafted the manuscript. All authors critically revised the manuscript for important intellectual content. PZ performed the statistical analysis. HY obtained the funding. All authors read and approved the final manuscript.

## Acknowledgements

Not applicable.

## Date availability

This research was conducted using the UK Biobank Resource under Application 61083. It has been stated in the Material Transfer Agreement of this project that data may be used solely by the Applicant PI and the related Applicant Researchers. However, data from the UK Biobank are open to researchers upon application to conduct health-related research in the public interest. Researcher can apply for the data through the link https://www.ukbiobank.ac.uk/enable-your-research/apply-for-access. The UK Biobank team will review the application.

## Code availability

All statistical analyses were conducted by the following software tool: Stata 15.1. All codes associated with the current submission is available, and can be requested by contacting the corresponding authors.

## Authors’ information

Not applicable.

---

**Supplementary figure1.**
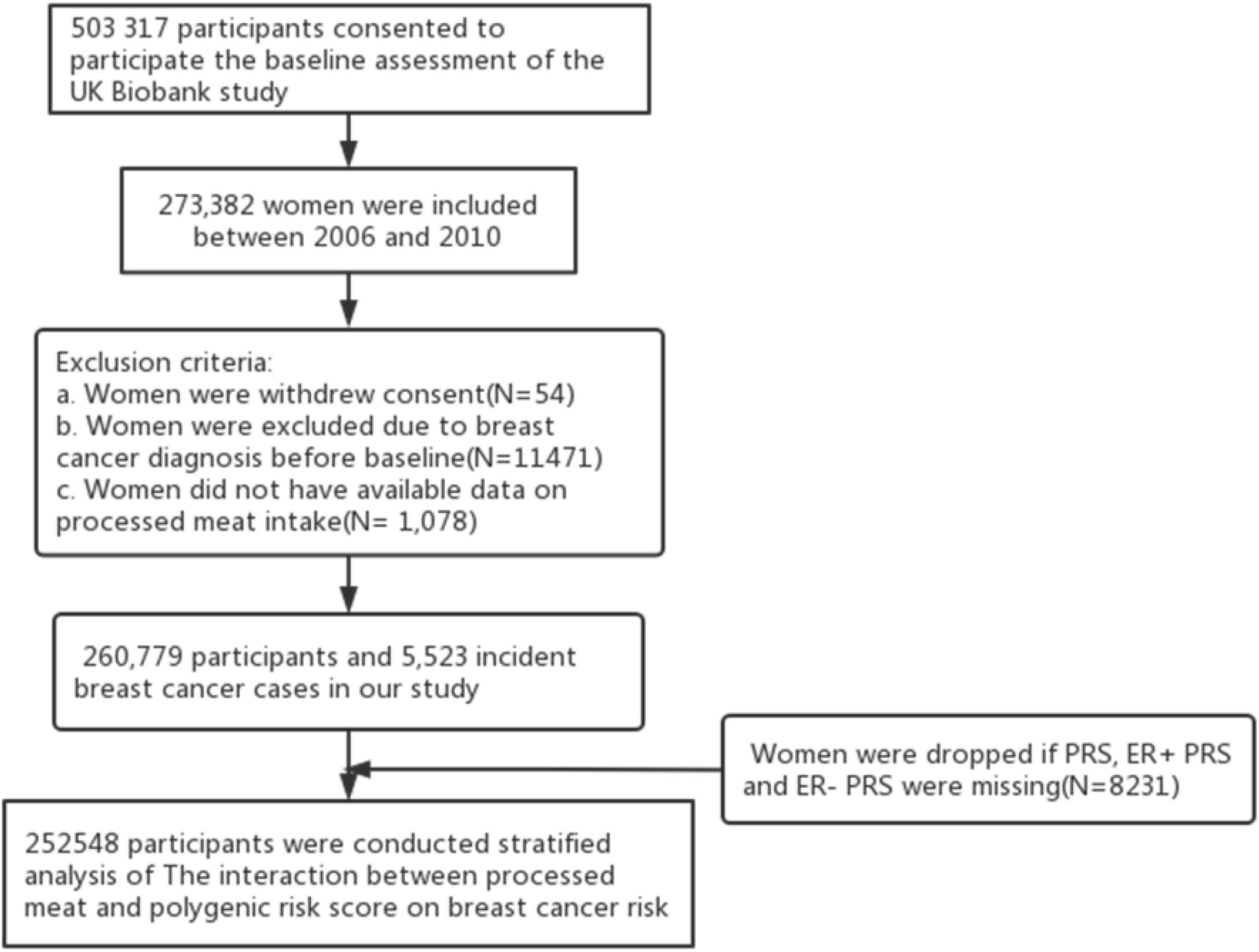
Flowchart of the study

## Notes

### Competing Interest Statement

The authors have declared no competing interest.

### Clinical Trial

This is not a clinical trial

### Author Declarations

The UK Biobank was approved by the National Information Governance Board for Health and Social Care and the National Health Service North West Centre for Research Ethics Committee (Ref: 11/NW/0382, 17 June 2011).

